# Variation in health visiting for the under 5s: A cross-sectional analysis of administrative data in England for 2018-2020

**DOI:** 10.1101/2024.02.22.24303170

**Authors:** Mengyun Liu, Jenny Woodman, Louise Mc Grath-Lone, Amanda Clery, Catherine Bunting, Samantha Bennett, Sally Kendall, Jennifer Kirman, Helen Weatherly, Jane Barlow, Helen Bedford, Katie Harron

## Abstract

**Introduction:** The health visiting service in England leads the delivery of the government’s Healthy Child Programme (HCP) for children under five. However, local authorities and their provider partners deliver this service differently in respond to the challenges of funding cuts, low numbers of health visitors and high levels of family needs.

**Objective:** We aimed to describe local authority variation in the delivery of health visiting services to children under 5 in England between 2018 and 2020.

**Methods:** We used nationally published statistics on mandated health visiting contacts, and administrative data from the Community Services Dataset (CSDS) on duration, location, and medium of delivery of contacts. We mapped the population coverage of mandated contacts (new birth visit, 6-8-week review, one-year review and 2-2 ½year review), and additional contacts, and described the frequency and characteristics of contacts across local authorities.

**Results:** According to the published statistics, 99% of eligible children received their new birth visit, 89% received the 6-8-week review and the one-year review, and 82% received the 2-2½-year review. There was substantial variation across local authorities: coverage for the 2-2½-year review ranged between 33%-97%. Based on CSDS, 80% of local authorities (n=46/57) delivered more additional contacts than mandated contacts: on average, 1.6 additional contacts (range: 0.1-8.5) were delivered for each mandated contact. There was also significant variation in the duration of contacts and the percentage of contacts delivered face-to-face and at home.

**Conclusions:** Our study demonstrates substantial variation in the delivery of health visiting services across England, particularly in the delivery of additional contacts. Further research is needed to explore the extent to which this trend has continued and the reasons for this variation. There is also a need to exploit this data to further explore the impact of these different models of service delivery on family outcomes.

## Introduction

The health visiting service in England leads the delivery of the government’s Healthy Child Programme (HCP) for children under five. The HCP comprises a universal preventative service and targeted support for families with higher need. [1-4] Health visiting teams are made up of health visitors (specialist community public health nurses), community staff nurses, nursery nurses, health care assistants, and other specialist health professionals. [2-4]

In England, health visiting teams are mandated to provide five universal health reviews: during the third trimester of pregnancy (health promoting visit), at child age 10-14 days (new birth visit), 6-8 weeks (6-8-week review), 12 months (one-year review), and 2-2½ years (2-2½ year review), each with a schedule of health promotion activities and with a review of health and development of the child within their family context (Supplementary material A, Appendix Table A1). [3] These mandated contacts provide an opportunity for health visiting teams to identify children and families in need of additional support, [3] which can comprise further additional contacts and/or referral of child and family to specialist services. [2, 3] Additional contacts might include support for the continuation of breastfeeding or advice on safer sleep, support for nutrition and accident prevention, or advice on managing minor illnesses. [3]

From 2015, the responsibility for commissioning public health services for children aged 0-5, including health visiting, was transferred from the NHS to local authorities. [5] Local authorities and their provider partners have faced funding cuts, low numbers of health visitors and high levels of family need, putting pressure on the service. [4, 6-8] Case studies and surveys have shown that local authorities have responded differently to these challenges, leading to additional variation in models of service delivery, priorities, and capacity. [2, 7, 9] To-date, there have been two analyses of routinely collected data on health visiting using the national individual-level administrative data on health visiting in England (the Community Services Dataset; CSDS). One study focused on variation in coverage and average number of health visiting contacts for children aged 2 years in 2018-9, by child-level characteristics (ethnicity, deprivation, child safeguarding flags). [10] This study found that found that children living in the most deprived neighbourhoods or with vulnerability markers (e.g. Looked After Child) were less likely to receive their 2–2½ year health and developmental review than other children but when all additional contacts were included, the pattern was reversed (deprivation) or disappeared (Looked After Children). There were no clear patterns by ethnicity and no analyses of local authority variation. [10] The second study reported similar results, based on a wider age range of children (birth to 3 years) and for two years of data (2018-20) and also reported high variation between local authorities in the proportion of mandated contacts conducted at home. [11] This current study adds to the evidence-base by providing a local authority view of health visiting delivery in terms of relative frequency of mandated to additional contacts from birth to age 5, a more detailed picture of medium and location and for the first time, contact duration and use of group contacts.

Our results highlight areas for improvement or investigation, can inform parameters for national guidance and local area decisions about service commissioning, including by benchmarking their own service. Our analyses also provide a reference point for international comparisons of child health services in the early years.

## Methods

### Data Sources

#### Health Visiting Service Delivery Metrics (Health Visiting metrics)

Health Visiting metrics are published by the Office of Health Inequalities and Disparities (OHID), who report local authority-level quarterly experimental statistics on health visiting service delivery, including the numbers of each mandated contact delivered and the number of eligible children. [12-14] These quarterly aggregate data are collected from commissioners of health visiting services in each local authority. [15] Health Visiting metrics provides the most accurate figures on the coverage of mandated contacts by local authorities, but does not include information on additional contacts or the format and duration of contacts. For each of the four postnatal mandated contacts, we extracted the number completed and the number of eligible children in each LA for two financial years from April 2018 to March 2020. As these data are experimental, we carried out additional validation and cleaning to correct reporting errors. [16]

#### Community Services Dataset (CSDS)

CSDS is an individual-level whole-population dataset established by NHS Digital in 2015, which comprises data on the utilization of all publicly funded community health services in England by individuals of all ages, including health visiting services. Relevant to this study, CSDS includes patient personal and demographic characteristics and health visiting activity. [14, 17]

We extracted data from CSDS for April 2018 to March 2020. We identified mandated health visiting contacts using the ‘Activity Type’ flag entered by the practitioner. [16] Where the Activity Type was missing (30% of records), we identified any health visiting contact that took place within a plausible time window for each mandated contact (Supplementary material A, Appendix Table A2). Additional contacts (i.e., health visiting contacts not defined as mandated) were identified for children up to 5 years old using the “Service or Team Type” variable indicating the health visiting service. We excluded contacts that were labelled as “did not attend”.

CSDS provides detailed information on how contacts are delivered but previous studies show that data in CSDS is incomplete and submitted inconsistently over time in some local authorities. [18] We assessed the completeness of CSDS at local authority-quarter level by comparing it to the Health Visiting metrics to create a “research-ready” dataset. [16] There were 149 local authorities (health visiting in City of London is delivered by Hackney and Isles of Scilly by Cornwall), each contributing up to 8 quarters of data, resulting in a maximum of 1,192 local authority-quarters possible for analysis. There were 57 local authorities (contributing 164 quarterly data points) that had high correlation with the Health Visiting metrics and were included in the “research-ready” dataset for analysis (Supplementary material A, Appendix Table A3). This means that we have ‘snap shot’ data by quarter for local authorities (Supplementary material A, Appendix Figure A1). The local authorities included in the “research-ready” dataset represent the distribution of regions, urban/rural status, and deprivation quintiles of England (Supplementary material A, Appendix Table A4). Further data cleaning is described in Supplementary material B.

### Indicators of health visiting delivery

We derived a set of local-authority level indicators for each type of health visiting contact (Supplementary material A, Appendix Table A5) and described the median and range of these indicators across local authorities. For local authorities with multiple quarters included in the “research-ready” dataset, we pooled data across quarters.

### Analysis

The geographic distribution of coverage of mandated contacts was mapped, and box plots were used to visualise variation in the medium (face-to-face or telephone) and locations (at home, health and social settings, or children’s centre) of health visiting delivery for both mandated and additional contacts across local authorities. All counts of individuals were rounded to the nearest 5 to reduce the risk of identifying individuals. [19]

## Results

All counts of contacts from CSDS have been rounded to the nearest 5 to comply with NHS statistical disclosure rules for subnational data.

### Study sample

There were 4,172,835 mandated contacts delivered to children aged 0-5 years between April 2018 and March 2020 in 149 local authorities, recorded in the Health Visiting metrics (Supplementary material A, Appendix Table A6). In the “research-ready” CSDS data for the same period, we included 1,779,155 contacts from 57 local authorities (Supplementary material Appendix A, Table A6).

### Coverage of mandated contacts (Health Visiting Metrics)

Coverage of the new birth visit was highest (median coverage of 98.5%, range: 89.9%-100.0%), followed by the 6-8-week review (median: 88.5%, range: 17.8%-100.0%) and one-year review (median: 89.0%, range: 19.9%-98.9%; Figure 1). The 2-2½-year review had a relatively lower coverage (median: 81.5%, range: 33.0%-97.4%). For 29 (19.5%) local authorities, >20% of eligible children did not receive the 6-8-week review. For 26 (17.4%) local authorities, >20% of eligible children were not recorded as receiving the one-year review. For 68 (45.6%) local authorities, >20% of eligible children were not recorded as receiving the 2-2½-year review and in 5 local authorities, >50% of eligible children were not recorded as having the 2-2½-year review.

**Figure 1.**
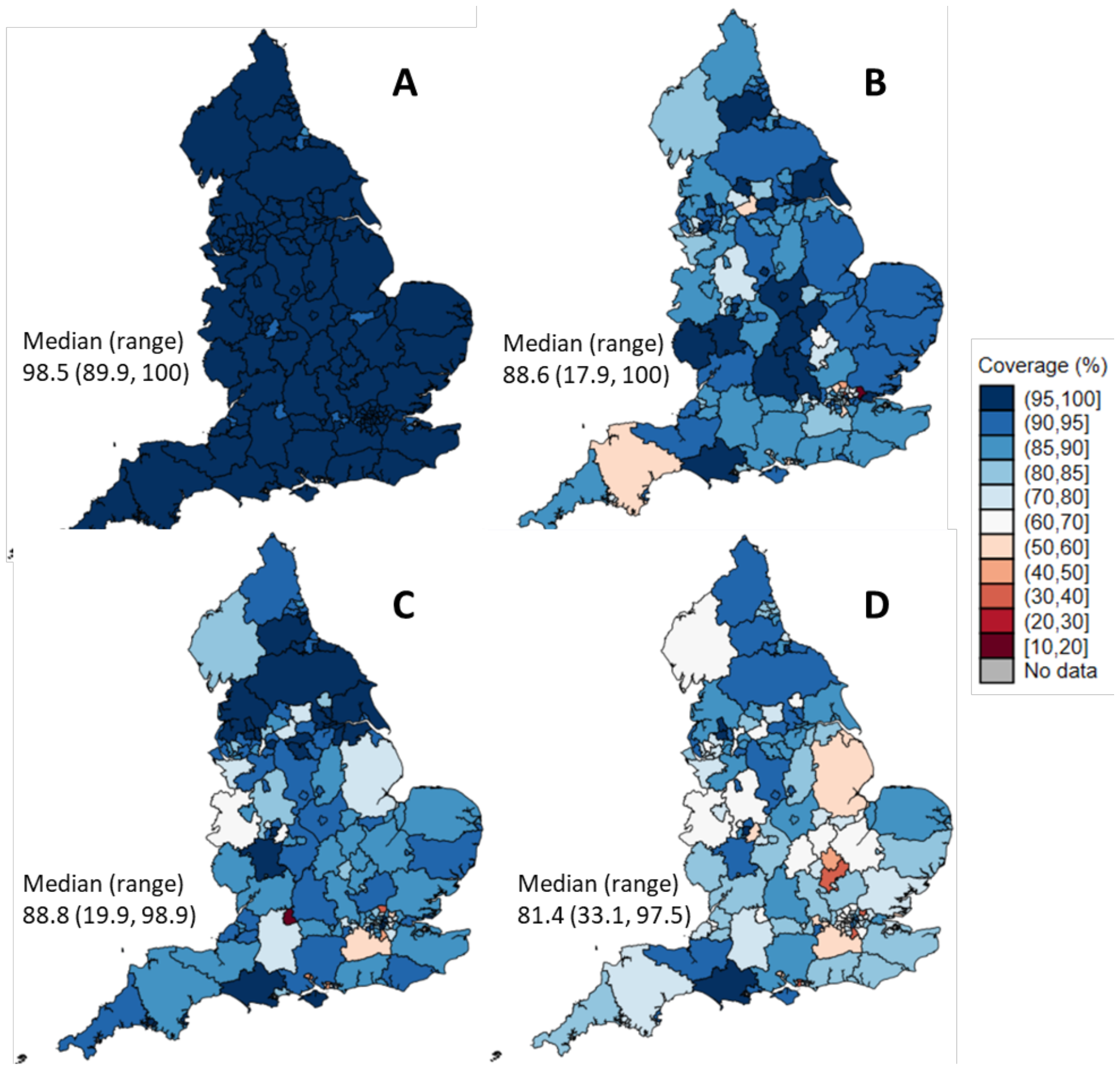
Coverage of mandated health visiting contacts in England based on Health Visiting Metrics, 2018/19-2019/20. A: coverage of new birth visit; B: coverage of 6-8-week review; C: coverage of one-year review; D: coverage of 2-2½-year review.

### Additional contacts (research-ready CSDS data)

Most local authorities (n=46, 80%) reported conducting more additional contacts than mandated contacts, with a median of 1.6 additional contacts delivered per mandated contact, and wide variation across local authorities (range 0.1-8.5). The number of additional contacts per mandated contact varied between 0.6-2.8 in 80% of local authorities, indicating that this variation was not due to a small number of outlying local authorities. Local authorities that delivered more health visiting contacts tended to conduct more additional contacts per mandated contact (Supplementary material A, Appendix Figure A2).

On average, 61.1% of children who received a health visiting contact received an additional contact within the year-quarter, but there was high variation across local authorities (range: 3.8%-94.1%). The large variation was not due to extreme values: in 80% of local authorities, the percentage of children who received a health visiting contact who received an additional contact within the year-quarter was between 27.2%-75.1%.

### Medium (research-ready CSDS data)

The majority (>96%) of mandated contacts were delivered face-to-face (Figure 2). Most additional contacts were also delivered face-to-face (median 82.7%, range: 47.6%-99.5%). About half of local authorities (n=23, 44.2%) offered >20% of additional contacts virtually. Phone calls were also used to deliver additional contacts: 34 local authorities (65%) delivered >10% of additional contacts through phone calls. Other mediums, including telemedicine, email, and messages, were rarely used for mandated or additional contacts, with <3% delivered through these mediums.

**Figure 2:**
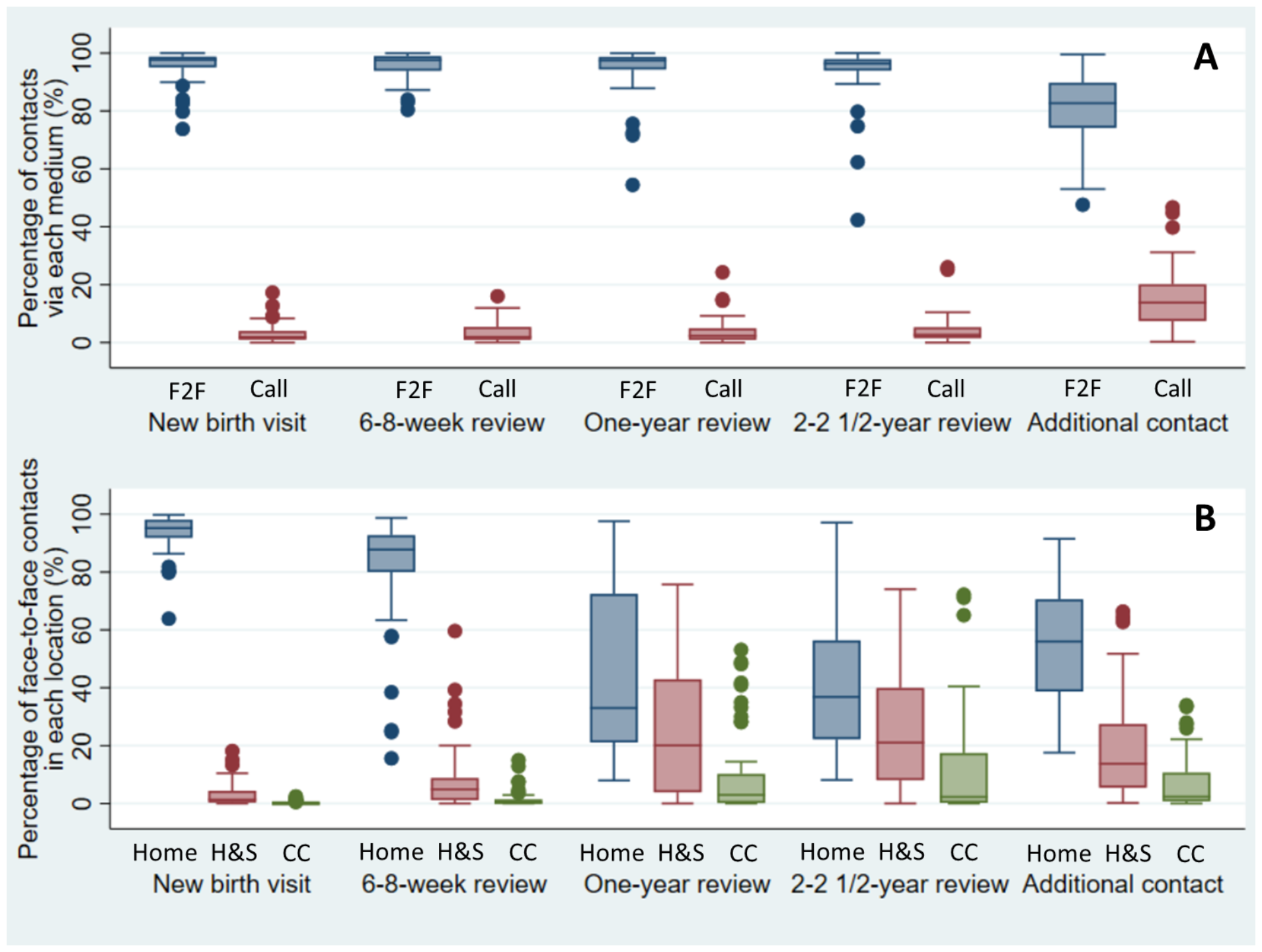
Variation in delivery of health visiting contacts based on research-ready CSDS data. A: Medium of delivery. B: Locations of face-to-face contacts. N=52 and N=49 local authorities with information about medium and location, respectively. F2F: face-to-face contacts; Call: phone call contacts; H&S: healthcare and social setting; CC: children’s centre

### Location (research-ready CSDS data)

New birth visits and 6-8-week reviews predominantly took place in the home setting, but there was substantial variation in the location of one-year reviews, 2-2½-year reviews and additional face-to-face contacts (Figure 3).

### Duration (research-ready CSDS data)

Mandated contacts were typically longer than additional contacts (Table 1), but high variation existed across local authorities. Face-to-face contacts were longer than phone call contacts. In more than half of the local authorities with available data on duration, 65% of new birth visits were at least an hour long, but most local authorities delivered more than half of other mandated contacts in 30-59 minutes. A considerable percentage of mandated contacts were recorded as lasting under 30 minutes (median 4.6%-7.9%) or 30-44 minutes long (median 12.3%-29.6%). Additional contacts were generally short: on average, 36.2% of additional contacts were completed in under 30 minutes. The duration of both mandated and additional contacts showed large variations across local authorities.

**Table 1.**
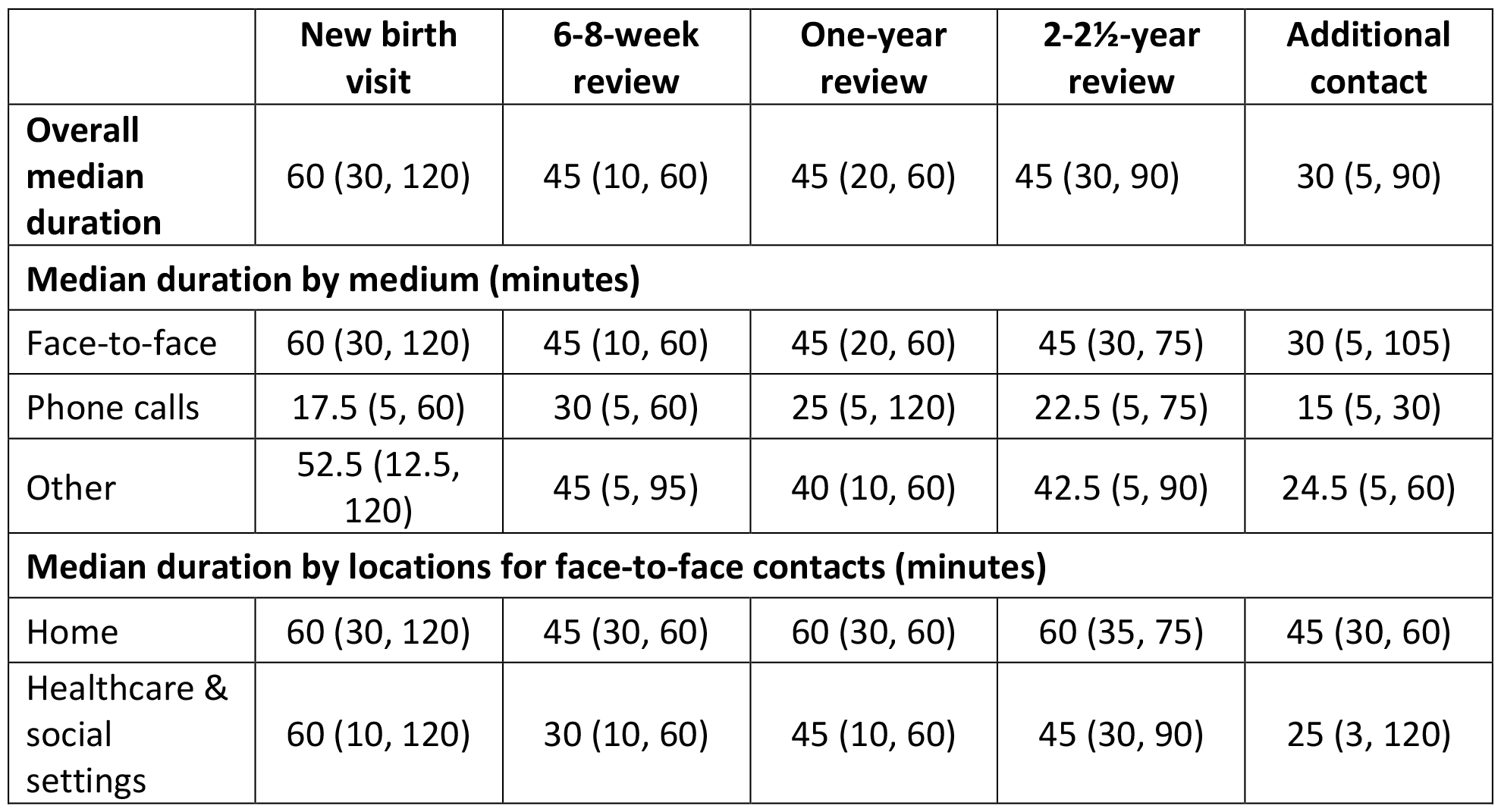
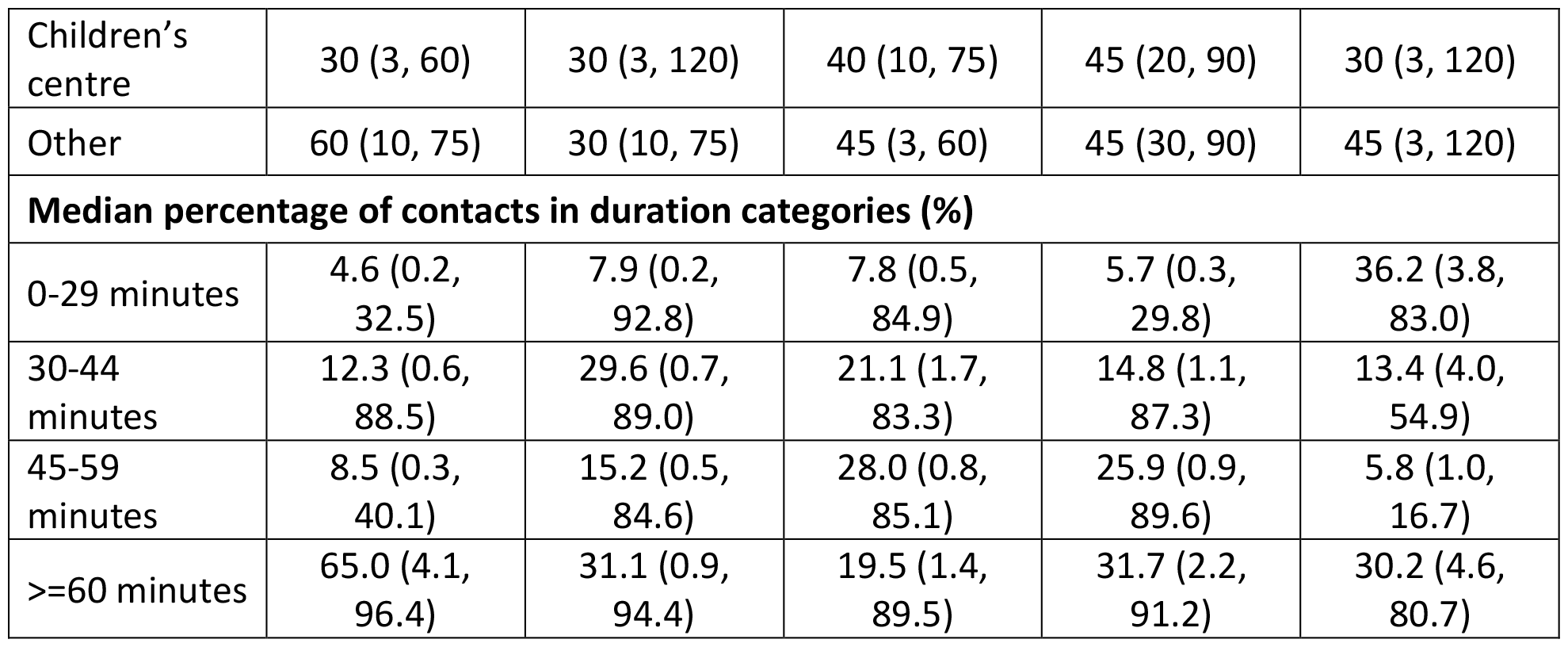
Duration of health visiting contacts in minutes, median duration or median percentage (range across local authorities). N=50 local authorities with data on duration

### Use of group sessions (research-ready CSDS data)

Group sessions were rarely used for mandated contacts. Of the 48 local authorities with information on group sessions, only five (10%) local authorities delivered >2% of new birth visits, 6-8-week reviews, and one-year reviews in group sessions. The use of group sessions also varied by local authority and type of contact.

## Discussion

### Key Findings

Between 2018-2020, the new birth visit was delivered with almost universal coverage but variation across England existed in the coverage of the 6-8-week, one-year, and 2-2½-year reviews. The high coverage of new birth visits and lower coverage of 2-2½-year review are consistent with previous studies using CSDS. [10, 11] However, our results also demonstrate that much of the reported activity was being delivered outside of mandated contacts, with 80% of local authorities delivering more additional than mandated contacts. The duration and way in which these additional contacts are delivered, for example by telephone or face-to-face, also varied across areas, which has not previously been described.

### Implications

There have been many years of austerity, cuts and a depleted workforce: since 2015 the real term value of the public health grant (from which health visiting is commissioned) has fallen by 26% [20] and the health visiting workforce has decreased by 37% from 11,193 in 2015 to 7,030 in 2022. [21] Despite this, health visiting teams during 2018-2020 were reaching nearly all babies and most children face-to-face, and conducting over one and a half times the number of additional contacts relative to mandated contacts. This represents a significant public health infrastructure to support the health and development of babies and children and the wellbeing of their families in the critical period before school. Our study highlights the importance of taking into account additional contacts when measuring health visiting activity. Additional contacts, to address identified need, were the most frequent type of contact (though shorter and more likely to be on the phone than the mandated reviews) in our analysis. This is consistent with other evidence about significant health visiting activity outside the mandated contacts in England, including a birth cohort study (of children born in 2020) which reported that 27% of 8628 families had four or more contacts with the health visiting team in the first 9 months of their child’s life. [22] However, there was substantial variation in the frequency, duration and medium of additional contacts, which is unlikely to be completely explained by differences in underlying need of the families across areas. Other factors that may have contributed to this variation include local service specifications and service priority, budgets, personnel capacity, and the availability of other local services. [23] Other evidence suggests that despite substantial ‘additional’ contacts, not all identified need can be followed up by the health visiting team: in a survey of 1186 health visitors, 45% reported that mandated contacts were prioritised over targeted or specialist support and 79% said that the service lacked capacity to offer a package of support to all children with identified needs. [24] Further qualitative work with parents and professionals is needed to understand the experiences and perceived purpose and impact of different types of additional contacts, as well as why models of additional contact are so different across areas.

Our results can inform conversations at the national and local level about the parameters of a ‘good’ health visiting service during a time when early years is a priority area, with targeted funding and innovation (e.g., Start for Life and Family Hubs [25]). For example, our findings raise a question about whether a mandated review can be adequately conducted in less than 45 minutes, given that it should include a direct observation of the child and significant health promotion activity. In this pre-Covid data we found that a considerable percentage of mandated contacts were recorded as lasing under 30 minutes (median 5%-8%, Table 1) or 30 to 44 (median 12%-30%) minutes. Providers, commissioners and national policy teams should consider how far features of delivery such as <30 minute or <45 minute mandated contacts are consistent with the way health visiting is theorised to work. In other words, is there a minimum time feasibly needed for health visitors or members of their team to build relationships with parents (including those with a mistrust of services), work in partnership with families, undertake health promotion work and conduct a holistic needs assessment of the whole family and family environment? It is possible that some features of service delivery, such as the shorter mandated contacts, are a service response to resource pressure which have the unintended consequence of undermining the mechanisms by which health visiting is thought to work and influence family outcomes. Next steps in building the evidence-base for national and local decision makers is to understand local authority variation in delivery to different types of families, including those living in deprivation or with other indicators of vulnerability, and the impact of decisions about service delivery on outcomes. [26, 27] As our study involved the use of data that was collected in the pre-pandemic era, we also now need to investigate how far the service delivery has ‘bounced back’ or has been permanently changed following the partial stop of health visiting services and widespread redeployment of staff and a change to virtual contacts. [28-31]

### Strengths and limitations

A strength of our study is that we used a subset of nationally representative data with high levels of completeness to monitor and measure the coverage, medium, location, duration of contacts, the delivery of additional contacts and use of group sessions. For estimates of coverage of mandated visits, we used aggregate Health Visiting metrics that cover every local authority in England. For the other characteristics of health visiting service delivery, we only included local authorities that correlated with health visiting activity recorded in the Health Visiting metrics (research-ready CSDS data). This means that misclassification of non-health visiting activity as mandated contacts for records with missing values on Activity Type is likely to be marginal.

Data quality remains an issue. First, we may not have identified all health visiting contacts in the CSDS, due to missing data in the “Service or Team Type” variable. This was likely a bigger issue for additional contacts, because we had no alternative method of identifying these (unlike mandated contacts for which we could use the “Activity Type” variable). [16] However, our study actually found a higher number of additional contacts than reported by other analyses of CSDS, which found 0.3 additional contacts per mandated contact for children between April 2019-March 2020 (compared to 1.6 in our study). [32] This may be explained by different ways of cleaning and curating the data: our research-ready data described health visiting on a subset of local authorities with more complete health visiting data, which may be associated with higher health visiting activity. There might also be misclassification between mandated and additional contacts: some local authorities offer more than five mandated contacts to every child and these extra “mandated contacts” will be identified as additional contacts in our analysis. Results about the use of additional contacts thus need to be interpreted with caution.

Results about the shortest mandated contacts should also be treated with caution (median 5%-8% of mandated contacts were recorded as under 30 minutes, Table 1). It is possible that some of these contacts were flagged as mandated contacts but were in fact initial conversations to schedule or prepare for a longer contact, which could have taken place shortly afterwards and covered some or all of the relevant mandated content. There is some evidence to support this hypothesis from our unpublished longitudinal analysis for a related health visiting project: [26] we found that 24% of children had an additional health visiting contact within two weeks of their new birth visit, but 41% of children with a new birth visit under 30 minutes had an additional contact within two weeks. Although the new birth visits under 30 minutes duration were more likely to be followed up within two weeks, most were not.

Second, we can only report on local authorities included in the research-ready dataset and we cannot assume that these findings are generalisable to local authorities with incomplete CSDS data. [10] The distribution of regions, urban/rural status, and deprivation quintiles in our analysis is representative to that in England (Supplementary material A, Appendix Table A4). Our findings are largely based on a few months of data within each area, as more than half of local authorities (n=31, 54.4%) included in the research-ready dataset only contributed one or two year-quarters. The sample selection largely depended on good reporting of data, which might be associated with good services. However, the coverage of mandated contacts is similar between the research-ready datasets and the whole dataset (Supplementary material A, Appendix Figure A3) and the median duration of 45-60 minutes for mandated contacts and 30 minutes for additional contacts is consistent with the average duration of 42 minutes pooling mandated and additional contacts together reported by health-visiting providers in a survey. [33]

Third, we were unable to conduct longitudinal analyses in this research-ready dataset, as a result of using the quarterly Health Visiting metrics as a reference to assess the completeness of CSDS (Supplementary material A, Appendix Figure A1). This means we were unable to describe the average number of contacts per child or patterns of contacts over early childhood.

Finally, as this study describes different aspects of service delivery independently, the association between them is not clear. For example, we do not yet know whether local authorities with a high coverage of mandated contacts had more additional contacts per mandated contact, or whether those who offered more additional contact per mandated contact had generally shorter contacts. Further exploration of models of service delivery combining different aspects together will help draw a more comprehensive picture of health visiting service delivery in England.

## Conclusion

This study demonstrates substantial variation in the delivery of health visiting across England, with potential unmet need in some local authorities. Further work is needed to explore whether this trend has continued in more recent years, and if so, the reasons for (for example, local specification and staff capacity) and determinants of (for example, urban/rural status and deprivation) this variation; it also provides a natural opportunity to exploit this variation to understand the impact of different models of service delivery on family outcomes.

This national picture of health visiting should be used by local authorities to review their own practice based on data in local systems, compare their practice with the national statistics and identify areas for improvement. [27] Such comparisons would generate hypotheses about the causes of outliers in terms of whether this is due to commissioning decisions or to the level of need and availability of other services in the area.

We demonstrate that with careful validation and cleaning, administrative data can be used to create a national picture of health visiting that can be used for research, monitoring and evaluation. However, we also highlight the need for data reporting and transfer systems to be strengthened in order to improve completeness and representativeness of these data to support future analyses.

## Acknowledgements and funding

This study/project is funded by the NIHR Public Health Research Programme (NIHR129901). This research was supported in part by the NIHR Great Ormond Street Hospital Biomedical Research Centre. This research benefits from and contributes to the NIHR Children and Families Policy Research Unit but was not commissioned through this Policy Research Unit (PR-PRU-1217-21301).

The views expressed are those of the author(s) and not necessarily those of the NIHR or the Department of Health and Social Care.

We thank study Steering Committee membership: Dr Louise Marryat, Donjeta Baliu, Dr Annie Herbert, Dr Toity Deave and Prof John Macleod. Thank you to Dr Cheryll Adams, CBE and Alison Morton for advice and input. We are grateful to all the Local Authority commissioners and NHS providers of health visiting services who met with us to sense-check our methods and interpretation of results.

## Ethics Statement

This study has been approved by University College London Institute of Education (UCL IOE) Research Ethics Committee (1531).

## Data Availability Statement

Access to the CSDS was approved and provided by NHS England (NIC-393510 and NIC-381972). Health Visiting Service Delivery Metrics data are published by the Office for Health Improvement and Disparities and are openly available: data for 2018/19 [12] can be found at https://www.gov.uk/government/statistics/health-visitor-service-delivery-metrics-2018-to-2019 and data for 2019/20 [13] can be found at https://www.gov.uk/government/statistics/health-visitor-service-delivery-metrics-experimental-statistics-2019-to-2020-annual-data.

### Supplementary Appendices

#### Supplementary Material A. Tables and Figures

Appendix Table A1. Focuses of each universal health review

Appendix Table A2. Coding of date-derived mandated health visiting contacts in the Community Service Dataset

Appendix Table A3. Local-authority-quarters included in the research-ready subset (with overall highly complete data on Health Visiting contacts in CSDS compared to the Health Visiting metrics)

Appendix Figure A1. Illustration of the selection of research ready dataset in an imaginary local authority

Appendix Table A4. The distribution of regions, urban/rural status, deprivation quintiles in local authorities included and not included in research ready dataset, N (%)

Appendix Table A5. Calculation of local-authority-quarter level indicator

Appendix Table A6. Characteristics of study sample in the Community Services Dataset “research-ready data”

Appendix Figure A2. Scatter plots showing the pattern of the number of additional contacts per mandated contact with the size of local authority, grouped by provider type (N=55)

Appendix Figure A3. Distribution of coverage of mandated contacts at the local-authority level, comparing “research-ready” dataset and the whole dataset, April 2018-March 2020

#### Supplementary Material B. Data cleaning and management process

Appendix Table B1. Missingness in variables relevant to Health Visiting service delivery in CSDS for research-ready dataset

Appendix Table B2. Recategorization of the medium and location of contacts

