## Appendix for "Variation in health visiting for the under 5s: A cross-sectional analysis of administrative data in England for 2018-2020"

### Supplementary Material A. Tables and Figures

**Appendix Table A1. Focuses of each universal health review**

| Health review | Age window | Focuses |
|---|---|---|
| Antenatal review | 28 weeks pregnancy | Breastfeeding, safer sleep, smoke free pregnancy, immunisation status, maternal and partner mental health |
| New birth review | 10-14 days after birth | Breastfeeding support, safer sleep, transition to parenthood, parent-child interaction, smoke free home |
| 6-8-week review | 6-8 weeks old | Breastfeeding support, family mental health, immunisation status, safer sleep, communication and interaction |
| One-year review | 9-12 months old | Immunisation status, nutrition, safer sleep, oral health, accident prevention, physical activity, speech, language, communication |
| 2-2½-year review | 2-2½ years old | Immunisation status, physical activity, nutrition, oral health, accident prevention, school readiness, speech, language, communication |

**Appendix Table A2. Coding of date-derived mandated health visiting contacts in the Community Service Dataset**

| Variable name | Relevant time period | Mandated contact |
|---|---|---|
| ContactBetween8_14Days_Flag | 8-14 days | New birth visit |
| ContactBetween15_30Days_Flag | 2-4 weeks | New birth visit |
| ContactBetween42_56Days_Flag | 6-8 weeks | 6-8-week review |
| ContactBetween42_63Days_Flag | 6-9 weeks | 6-8-week review |
| ContactBetween270_366Days_Flag | 9-12 months | 1-year review |
| ContactBetween270_457Days_Flag | 9-15 months | 1-year review |
| ContactBetween691_914Days_Flag | 1y 11 months-2y 6 months | 2-2½-year review |

**Appendix Table A3. Local-authority-quarters included in the research-ready subset (with overall highly complete data on Health Visiting contacts in CSDS compared to the Health Visiting metrics)**

| Number of quarters a Local Authority contributed | | | | Number of Local Authorities contributing to each quarter | | | |
|---|---|---|---|---|---|---|---|
| **Number of quarters** | **Number of LAs** | **Percent (%)** | **Cum percent (%)** | **Quarter** | **Number of LAs** | **Percent (%)** | **Cum percent (%)** |
| 1 | 15 | 26.3 | 26.3 | 2018/19 Q1 | 15 | 9.2 | 9.1 |
| 2 | 16 | 28.1 | 54.4 | 2018/19 Q2 | 16 | 9.8 | 18.9 |
| 3 | 11 | 19.3 | 73.7 | 2018/19 Q3 | 22 | 13.4 | 32.3 |
| 4 | 4 | 7.0 | 80.7 | 2018/19 Q4 | 22 | 13.4 | 45.7 |
| 5 | 3 | 5.3 | 86.0 | 2019/20 Q1 | 24 | 14.6 | 60.4 |
| 6 | 5 | 8.8 | 94.7 | 2019/20 Q2 | 22 | 13.4 | 73.8 |
| 7 | 1 | 1.8 | 96.5 | 2019/20 Q3 | 23 | 14.0 | 87.8 |
| 8 | 2 | 3.5 | 100 | 2019/20 Q4 | 20 | 12.2 | 100 |
| Total | 57 | 100 | | Total | 164 | 100 | |

Note: There are a total of 149 local authorities in the analysis as City of London is combined with Hackney and Isles of Scilly is combined with Cornwall. LA: local authority. Cum percent: cumulative percentage.

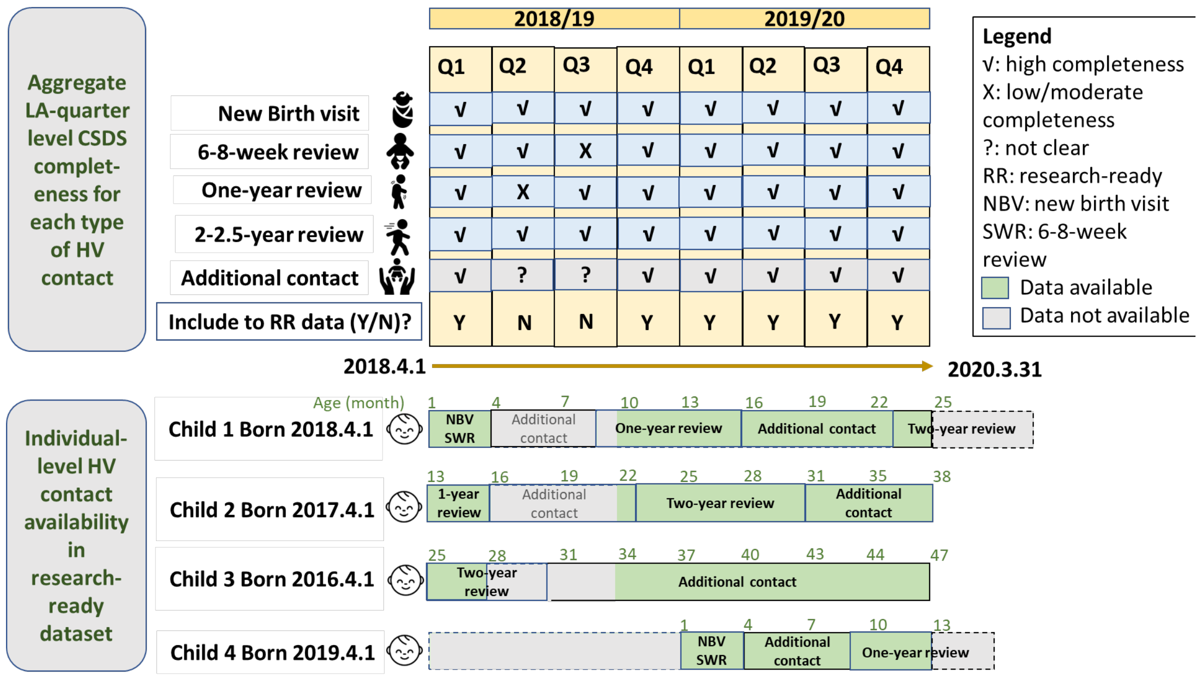


**Appendix Figure A1. Illustration of the selection of research ready dataset in an imaginary local authority**

**Appendix Table A4. The distribution of regions, urban/rural status, deprivation quintiles in local authorities included and not included in research ready dataset, N (%)**

| Characteristics | 149 local authorities in England | 57 local authorities included in research-ready CSDS dataset | 92 local authorities NOT included in research-ready CSDS dataset | P values comparing local authorities that are included with not included in the research-ready dataset |
|---|---|---|---|---|
| **Region** | | | | |
| East Midlands | 9 (6.0) | 4 (7.0) | 5 (5.4) | 0.79 |
| East of England | 12 (8.1) | 4 (7.0) | 8 (8.7) | |
| London | 32 (21.5) | 8 (14.0) | 24 (26.1) | |
| North East | 12 (8.1) | 6 (10.5) | 6 (6.5) | |
| North West | 23 (15.4) | 8 (14.0) | 15 (16.3) | |
| South East | 18 (12.1) | 8 (14.0) | 10 (10.9) | |
| South West | 14 (9.4) | 7 (7.6) | 7 (12.3) | |
| West Midlands | 14 (9.4) | 6 (10.5) | 8 (8.7) | |
| Yorkshire and The Humber | 15 (10.1) | 6 (10.5) | 9 (9.8) | |
| **Urban/rural status** | | | | |
| Predominantly rural | 20 (13.4) | 9 (15.8) | 11 (12.0) | 0.79 |
| Predominantly urban | 108 (72.5) | 40 (70.2) | 68 (73.9) | |
| Urban with significant rural | 21 (14.1) | 8 (14.0) | 13 (14.1) | |
| **Index of multiple deprivation (IMD) quintiles** | | | | |
| Lowest quintile | 30 (20.4) | 12 (21.1) | 18 (20) | 0.68 |
| 2 | 29 (19.7) | 12 (21.1) | 17 (18.9) | |
| 3 | 30 (20.4) | 10 (17.5) | 20 (22.2) | |
| 4 | 29 (19.7) | 9 (15.8) | 20 (22.2) | |
| Highest quintile | 29 (19.7) | 14 (24.6) | 15 (16.7) | |
| **Income Deprivation Affecting Children Index (IDACI) quintiles** | | | | |
| Lowest quintile | 30 (20.1) | 12 (21.1) | 18 (19.6) | 0.92 |
| 2 | 30 (20.1) | 10 (17.5) | 20 (21.7) | |
| 3 | 30 (20.1) | 11 (19.3) | 19 (20.7) | |
| 4 | 30 (20.1) | 11 (19.3) | 19 (20.7) | |
| Highest quintile | 29 (19.5) | 13 (22.8) | 16 (17.4) | |

Note: There are a total of 149 local authorities in the analysis as City of London is combined with Hackney and Isles of Scilly is combined with Cornwall.

**Appendix Table A5. Calculation of local-authority-quarter level indicator**

| Indicator | Calculation | Data Source |
|---|---|---|
| **Coverage** | | |
| Coverage of mandated contacts | The number of eligible children who received a mandated contact/the number of children eligible for this mandated contact.<br><br>For example, for new birth visit, the coverage is calculated as 'the number of new birth visits'/ 'the number of children eligible for new birth visits'. This indicator is calculated separately for new birth visit, 6-8-week review, one-year review and 2-2½-year review. | Health Visiting metrics |
| Percentage of children with additional contacts | The number of children who received an additional contact/the number of children who received any Health Visiting contact (either mandated or additional). This indicator was calculated within each quarter-year. | CSDS |
| **Number of additional contacts per mandated contact** | | |
| Number of additional contacts per mandated contact | The number of additional contacts/the number of mandated contacts | CSDS |
| **Medium** | | |
| Percentage of contacts that are face-to-face | The number of face-to-face contacts/the total number of Health Visiting contacts* | CSDS |
| Percentage of contacts that are phone calls | The number of phone call contacts/the total number of Health Visiting contacts* | CSDS |
| **Location** | | |
| Percentage of face-to-face contacts at home | The number of face-to-face contacts at home/the total number of face-to-face contacts* | CSDS |
| Percentage of face-to-face contacts at healthcare or social settings | The number of face-to-face contacts at healthcare or social settings/the total number of face-to-face contacts* | CSDS |
| Percentage of face-to-face contacts at children's centres | The number of face-to-face contacts at children's centre/the total number of face-to-face contacts* | CSDS |
| **Duration** | | |
| Median duration | The median duration among all contacts* | CSDS |
| % mandated contacts <30 min | The number of mandated contacts in each duration category/the total number of mandated contacts* | CSDS |
| % mandated contacts 31-45 min | | CSDS |
| % mandated contacts 46-60 min | | CSDS |
| % mandated contacts >61 min | | CSDS |
| % additional contacts <15 min | The number of additional contacts in each duration category/the total number of additional contacts | CSDS |
| % additional contacts 16-30 min | | CSDS |
| % additional contacts 31-45 min | | CSDS |
| % additional contacts 46-60 min | | CSDS |
| % additional contacts >61 min | | CSDS |
| **Use of group session** | | |

| | | |
|---|---|---|
| Percentage of contacts that are delivered in a group session | The number of group session contacts/the total number of contacts* | CSDS |

* This indicator is calculated separately for new birth visit, 6-8-week review, one-year review, 2-2½-year review, and additional contacts.

**Appendix Table A6. Characteristics of study sample in the Community Services Dataset "research-ready data"**

| **Characteristic** | **CSDS "Research Ready Data"** | **Health Visiting metrics** |
|---|---|---|
| **Number of local authorities-quarters (local authorities)** | 164 (57) | 1192 (149) |
| **Average number of quarters per local authority (range)** | 2.9 (1, 8) | 8 (8, 8) |
| **Number of local authorities by main provider (%)** | | |
| NHS | 43 (75.4%) | - |
| Council | 6 (10.5%) | - |
| Others (e.g., private company) | 8 (14.0%) | - |
| **Number of contacts (%)** | 1,779,155 | 4,172,835 |
| New birth visits | 167,425 (9.4%) | 1,135,310 (27.2%) |
| 6-8-week-review | 157,130 (8.8%) | 1,006,025 (24.1%) |
| One-year review | 169,495 (9.4%) | 1,041,825 (25.0%) |
| 2-2½-year review | 165,680 (9.3%) | 989,680 (23.7%) |
| Additional contacts | 1,119,420 (62.9%) | - |

Note: There are a total of 149 local authorities in the analysis as City of London is combined with Hackney and Isles of Scilly is combined with Cornwall.

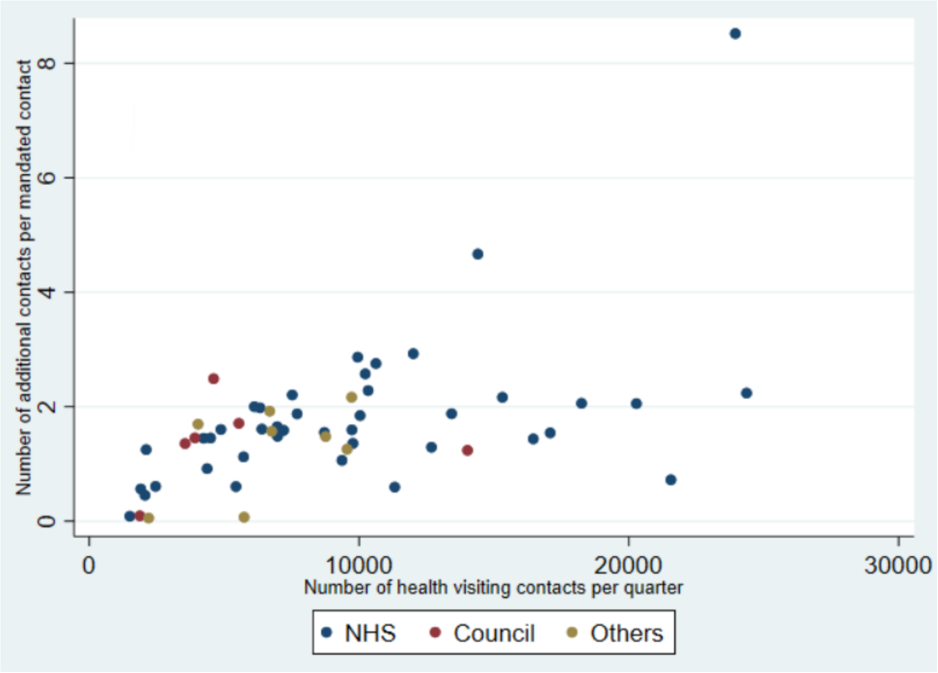


**Appendix Figure A2. Scatter plots showing the pattern of the number of additional contacts per mandated contact with the size of local authority, grouped by provider type (N=55)** Two local authorities with >30,000 health visiting contacts per quarter are not shown in this figure.

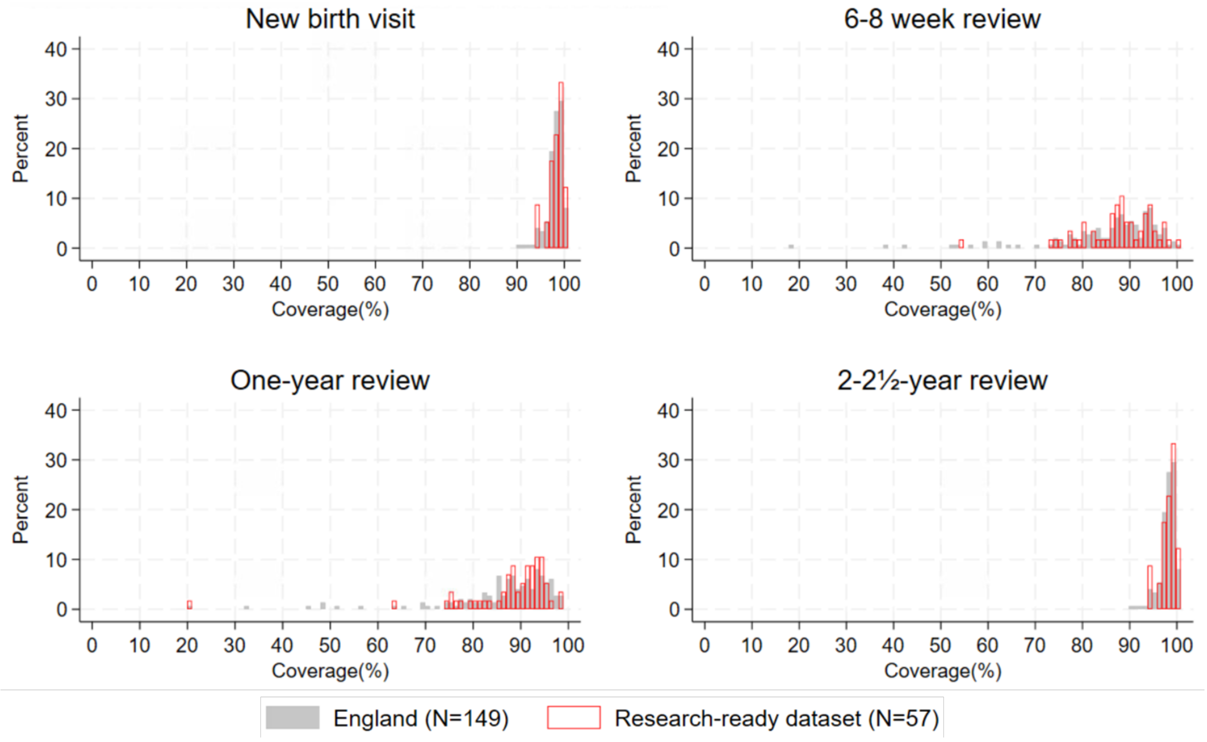


**Appendix Figure A3. Distribution of coverage of mandated contacts at the local-authority level, comparing "research-ready" dataset and the whole dataset, April 2018-March 2020**

**Supplementary Material B. Data cleaning and management process**

We checked the overall level of missing values for variables in Community Services Dataset (CSDS) that are relevant to health visiting delivery (Appendix Table B1). Locations were only applicable for face-to-face contacts. We include variables that are conceptually relevant and have acceptable level of missing values (<20%): medium, location, whether delivered in group setting, organisation identifier, and clinical duration. Some important variables were excluded due to high level of missing data, for example, staff type and occupation type.

**Appendix Table B1. Missingness in variables relevant to Health Visiting service delivery in CSDS for research-ready dataset (164 Local-authority-quarters from 57 local-authorities with *high completeness* of Health Visiting contacts for all four mandated contacts)**

| Variable | % contacts have missing values prior to cleaning | Comments |
|---|---|---|
| **Medium** (face-to-face, telephone, email, etc.) | 7.1% | Include |
| **Locations for face-to-face contacts** (home, health centre, child centre, others) | 12.8% | Include |
| **Group therapy** (yes/no) | 5.8% | Include |
| **Organisation identifier** | 0% | Include |
| **Clinical duration** (minutes) | 9.4% | Include |
| **Initial/follow-up contact** | 11.8% | Exclude: 1) most of the initial contacts are new birth visits and other mandated and additional contacts are follow-up contacts. 2) The "research ready dataset" is not longitudinal so for follow-up contacts, we may not include the initial contacts. make this variable not meaningful. |
| **Distance from home to contact location** | | Exclude: due to high levels of missing data |
| Home visit contacts | 82.0% | |
| In heath care and social settings | 68.3% | |
| In children's centres | 89.7% | |
| **Days referral to care contact** | 0% | Exclude: although no missing values, it is not clear what this variable means. |
| **Registration body** (general medical council, general dental council, health and care professions council, nursing and midwifery council, etc.) | 77.4% | Exclude: due to high levels of missing data |
| mandated contacts | 74.8% | Exclude: due to high levels of missing data |
| additional contacts | 78.9% | Exclude: due to high levels of missing data |
| **Staff type** (each type of therapist, health care worker or assistant, general medical practitioner, community midwife, health visitor, staff nurse, other nurses) | 79.2% | Exclude: due to high levels of missing data |
| new birth visit | 73.8% | Exclude: due to high levels of missing data |

| | | |
|---|---|---|
| 6-8 week review | 74.4% | Exclude: due to high levels of missing data |
| one year review | 78.6% | Exclude: due to high levels of missing data |
| two-year review | 81.5% | Exclude: due to high levels of missing data |
| additional contact | 80.4% | Exclude: due to high levels of missing data |
| **Occupation type** (admin staff, health care assistants & support staff, nursing & midwifery & Health Visiting staff, nursing & midwifery & Health Visiting learners, etc.) | 96.8% | Exclude: due to high levels of missing data |
| **Job role** (medical & dental, students, specialist nurse, staff nurse, community practitioner, etc.) | 77.9% | Exclude: due to high levels of missing data |
| mandated contacts | 75.5% | Exclude: due to high levels of missing data |
| additional contacts | 79.4% | Exclude: due to high levels of missing data |
| **Service/team type** (Health Visiting service, school nursing service, etc.) | 1.9% | Exclude: this variable was used to identify Health Visiting contacts |

We then cleaned and re-categorised the variables we included. For categorical variables, we checked errors. We speculate the true value where possible. For example, location variable was coded with organisation codes in some contacts, and we replaced the location variable with the corresponding location categories for these contacts. Where such speculation cannot be made, we set the errors to missing.

Categorical variables were then re-categorised into broader groups (Appendix Table B2). Provider organisations were grouped into three types: NHS providers, council providers, and other providers (including private organisations). Provider type was obtained by searching organisation identifier in CSDS on NHS Digital Organisation/Practitioner Search platform. The type of provider recorded for more than 80% of the Health Visiting contacts in a local authority was considered the main provider in this local authority. The medium of contact was grouped into face-to-face, phone calls, and others. We only looked at location of face-to-face contacts. Contact locations were grouped into home, health and social care settings, children's centre, and others.

**Appendix Table B2. Recategorization of the medium and location of contacts**

| **Categories in raw CSDS** | | **Categories used in analysis** | |
|---|---|---|---|
| **Code** | **Category** | **Code** | **Category** |
| **Medium** | | | |
| 01 | Face-to-face communication | 1 | Face-to-face contact |
| 02 | Telephone | 2 | Phone call |
| 03 | Telemedicine | 3 | Others |
| 04 | Talk type for a person unable to speak | | |
| 05 | Email | | |
| 06 | Short Message Service (SMS) – Text Messaging | | |
| 07 | Online Triage | | |
| 08 | Online Instant Messaging | | |
| 98 | Other (not listed) | | |
| **Location** | | | |

| | | | |
|---|---|---|---|
| A01-A04 | A01 patient's home; A02 carer's home; A03 patient's workplace; A04 other patient related location | 1 | Home |
| B01-B02 | B01 primary care health centre; B02 polyclinic | 2 | Health and social care settings |
| C01-C03 | C01 general medical practitioner practice; C02 dental practice; C03 ophthalmic medical practitioner premises | | |
| D01-D03 | D01 walk in centre; D02 out of hours centre | | |
| E01-E099 | E01 out-patient clinic; E02 ward; E03 day hospital; E04 emergency care department or minor injuries department; E99 other departments | | |
| H01 | day centre | | |
| J01 | resource centre | | |
| F01 | hospice | | |
| G01-G04 | G01 care home without nursing; G02 care home with nursing; G03 children's home; G04 integrated care home without nursing and care home with nursing | | |
| K01-K02 | K01 sure start children's centre; K02 child development centre | 3 | Children’s centre |
| L01-L99 | L01 school; L02 further education college; L03 university; L04 nursery premises; L05 other childcare premises; L06 training establishments; L99 other educational premises | 4 | Others |
| M01-M07 | M01 prison; M02 probation service premises; M03 police station / police custody suite; M04 young offender institution; M05 immigration removal centre; | | |
| N01-N03 | N01 street or other public open space; N02 other publicly accessible area or building; N03 voluntary or charitable agency premises | | |
| X01 | other locations not elsewhere classified | | |

For duration of contacts, we checked extreme values. Duration of zero minute was set to missing and durations that are longer than eight hours were set to eight hours. We used different duration categorisation for mandated contacts and additional contacts, as mandated contacts were generally longer than additional contacts. Mandated contacts are assigned into one of the following duration groups: 1-30 min, 31-45 min, 46-60 min, 61-75 min, and >75 min. Duration groupings used for additional contacts are: 1-15 min, 16-30 min, 31-45 min, 46-60 min, 61-75 min, and >75 min.

We further assessed missingness for these variables at the local-authority level. If a local authority had more than a quarter of data missing in a certain variable across all quarters within the dataset, we set the entire variable missing for this local authority as the quality of data is not reliable for this variable this local authority. This means that we may have lost some information on certain variables for some local authorities.
